# Original research: How accurate are digital symptom assessment apps for suggesting conditions and urgency advice?: a clinical vignettes comparison to GPs

**DOI:** 10.1101/2020.05.07.20093872

**Authors:** Stephen Gilbert, Alicia Mehl, Adel Baluch, Caoimhe Cawley, Jean Challiner, Hamish Fraser, Elizabeth Millen, Jan Multmeier, Fiona Pick, Claudia Richter, Ewelina Türk, Shubhanan Upadhyay, Vishaal Virani, Nicola Vona, Paul Wicks, Claire Novorol

## Abstract

**Objectives:** To compare breadth of condition coverage, accuracy of suggested conditions and appropriateness of urgency advice of 8 popular symptom assessment apps with each other and with 7 General Practitioners.

**Design:** Clinical vignettes study.

**Setting:** 200 clinical vignettes representing real-world scenarios in primary care.

**Intervention/comparator:** Condition coverage, suggested condition accuracy, and urgency advice performance was measured against the vignettes’ gold-standard diagnoses and triage level.

**Primary outcome measures:** Outcomes included (i) proportion of conditions “covered” by an app, i.e. not excluded because the patient was too young/old, pregnant, or comorbid, (ii) proportion of vignettes in which the correct primary diagnosis was amongst the top 3 conditions suggested, and, (iii) proportion of “safe” urgency level advice (i.e. at gold standard level, more conservative, or no more than one level less conservative).

**Results:** Condition-suggestion coverage was highly variable, with some apps not offering a suggestion for many users: in alphabetical order, Ada: 99.0%; Babylon: 51.5%; Buoy: 88.5%; K Health: 74.5%; Mediktor: 80.5%; Symptomate: 61.5%; Your.MD: 64.5%. The top-3 suggestion accuracy (M3) of GPs was on average 82.1±5.2%. For the apps it was – Ada: 70.5%; Babylon: 32.0%; Buoy: 43.0%; K Health: 36.0%; Mediktor: 36.0%; Symptomate: 27.5%; WebMD: 35.5%; Your.MD: 23.5%. Some apps exclude certain user groups (e.g. younger users) or certain conditions - for these apps condition-suggestion performance is generally greater with exclusion of these vignettes. For safe urgency advice, tested GPs had an average of 97.0±2.5%. For the vignettes with advice provided, only three apps had safety performance within 1 S.D. of the GPs (mean) - Ada: 97.0%; Babylon: 95.1%; Symptomate: 97.8%. One app had a safety performance within 2 S.D.s of GPs - Your.MD: 92.6%. Three apps had a safety performance outside 2 S.D.s of GPs - Buoy: 80.0% (*p*<0.001); K Health: 81.3% (*p*<0.001); Mediktor: 87.3% (*p*=1.3×10_-3_).

**Conclusions:** The utility of digital symptom assessment apps relies upon coverage, accuracy, and safety. While no digital tool outperformed GPs, some came close, and the nature of iterative improvements to software offers scalable improvements to care.

**Article Summary:** Strengths and limitations of this study

Strengths of the study include a large number of vignettes, peer-reviewed by independent and experienced primary care physicians to minimise bias.

Furthermore, GPs and apps were tested with vignettes in a manner that simulates real clinical consultations, based on mock telephone consultations, with detailed source data verification.

Vignette entry was conducted by professionals; a recent study found that laypeople are less good at entering vignettes for symptoms that they have never experienced.

Limitations include the lack of a rigorous and comprehensive selection process to choose the 8 apps and the lack of real patient experience assessment. Because software is constantly evolving, our findings cannot necessarily be generalized in the future. Future replication by independent researchers is needed.

## Introduction

Against the background of an aging population and rising pressure on medical services, the last decade has seen the internet replace general practitioners as the first port of call for health information. A 2010 survey of over 12,000 people from 12 countries reported that 75% of respondents search for health information online [1], with some two thirds of patients in 2017 reporting that they “google” their symptoms before going to the doctor’s office [2]. However, online search tools like Google or Bing were not intended to provide medical advice and risk offering irrelevant or misleading information [3]. One potential solution is dedicated symptom assessment applications (i.e. apps) [3–6], which use a structured interview or multiple-choice format to ask patients questions about their demographic, relevant medical history, symptoms, and presentation. In the first few screening questions some symptom assessment apps exclude patients from using the tool if they are too young, too old, are pregnant, or have certain comorbidities, limiting the “coverage” of the tool. Assuming the patient is not excluded, these software tools use a range of computational approaches to suggest one or more conditions that might explain the symptoms (e.g. common cold vs pneumonia). Many symptom assessment apps then suggest next steps that patients should take (levels of urgency advice, e.g. self-care at home vs seek urgent consultation), often along with evidence-based condition information for the user.

A recent systematic review of the literature identified that rigorous studies are required to show that these apps provide safe and reliable information [4] in the context for which they were designed and for which they have regulatory approval. Most previous studies considered only a single symptom assessment app, focused on specific (often specialty) conditions, had a small number of vignettes (<50), were relatively uncontrolled in the nature of the cases presented, and suffered a high degree of bias [4]. For example, a previous study examined the performance of the Mediktor app in the Emergency Department waiting room [7]. While this is a valid setting, most apps were designed and approved for use primarily at home and for newly presenting problems; accordingly, some 38.7% of patients had to be excluded. Few studies have systematically compared symptom assessment apps to one another in this context, which is particularly important as apps may increasingly be used to supplement or replace telephone triage [4]. This is particularly relevant in 2020 due to the COVID-19 pandemic - early in the spread of COVID-19, healthcare facilities risked being overwhelmed and furthering contagion, so communication strategies were needed to provide patients with advice without face-to-face contact [8,9].

In contrast to deploying apps in a heterogeneous real-world setting, where participants would not have the time to re-enter their symptoms multiple times, and may not receive a verifiable diagnosis, clinical vignettes studies allow direct comparison of inter-app and app-to-GP performance [10–12]. Clinical vignettes are created to represent patients, these are reviewed and then assigned gold-standard answers for main and differential diagnoses and for triage. The clinical vignettes are then used to test both apps and GPs. GPs are assessed through mock telephone consultations and apps through their normal question flow [3,6]. Clinical vignettes studies have the advantage of enabling direct GP-to-app comparison, allowing a wide range of case types to be explored, and are generalizable to “real life” situations, but are complementary to, not a replacement for, real-patient studies [4,10,12]. Seminal work at Harvard Medical School has established the value of such approaches but has not been updated recently [3,6].

The objective of the current study was to compare the coverage, suggested condition accuracy, and urgency advice accuracy of GPs and 8 popular symptom assessment apps: Ada, Babylon, Buoy, K Health, Mediktor, Symptomate, WebMD, and Your.MD.

## Methods

The process for clinical vignette creation, review, and testing of the GPs and the apps using the vignettes is shown in **Fig. 1**.

**Figure 1.**
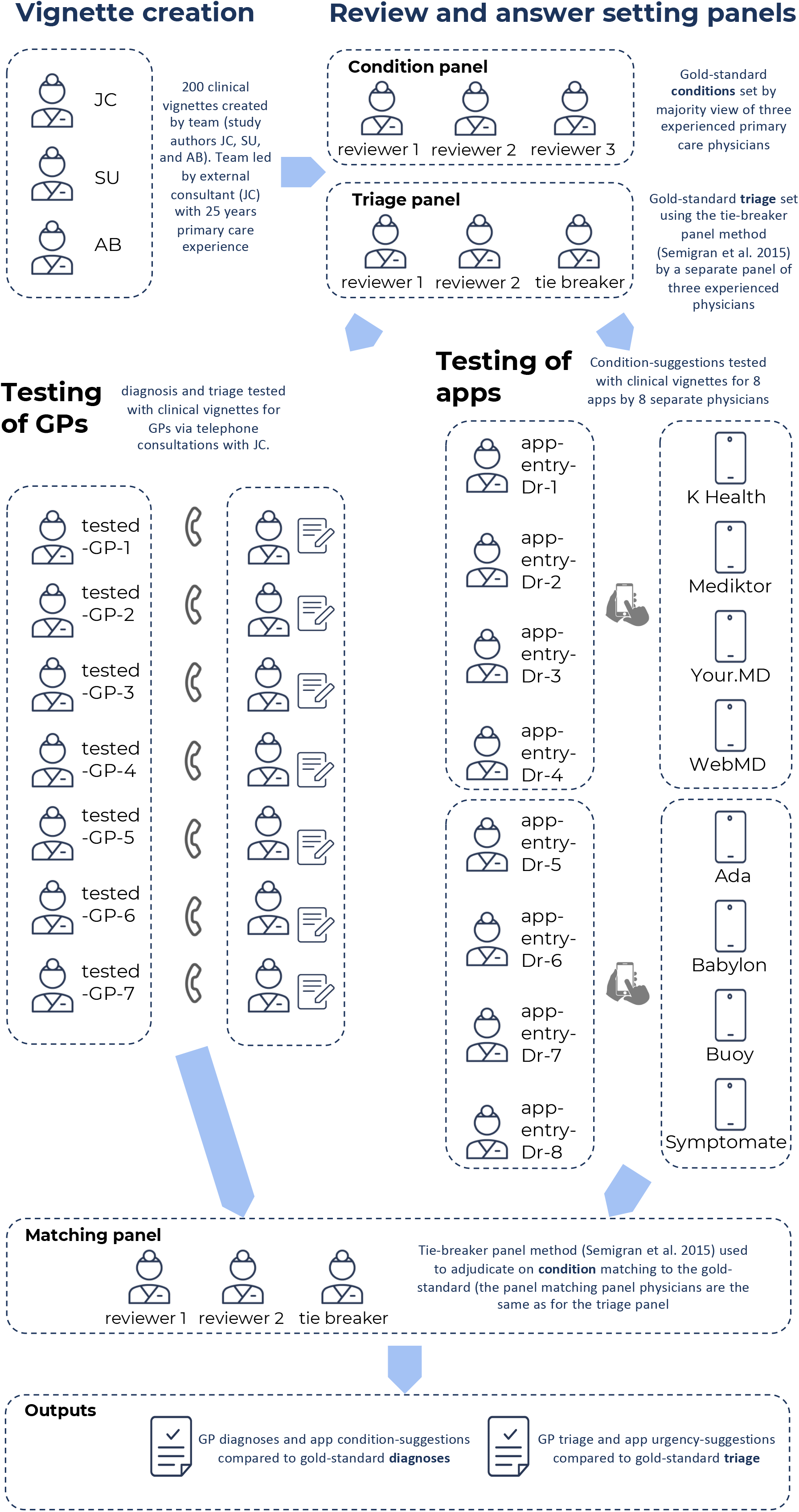
Overview of the study methodology including: (i) vignette creation; (ii) vignettes review and answer setting; (iii) testing of GPs; and, (iv) adjudication of matching of condition suggestions to the gold standard.

### Clinical vignette creation

An independent primary care clinical expert consultant (JC) was commissioned to lead the creation of 200 clinical vignettes: JC has over 25 years’ experience in general practice and emergency practice and has also had many years of experience in creating and customising algorithms for use in telephone triage and for internet-based self-assessment, including for *NHS Direct*. The vignette creation team also included two GPs (SU and AB - employees of Ada Health), each with over 5 years primary care and emergency department experience. SU and AB had worked for the Ada Health telehealth service *Dr Chat* but were not involved in the development of Ada’s medical intelligence. The vignettes were designed to include both common and less - common conditions relevant to primary care practice, and to include clinical presentations and conditions affecting all body systems. They were created to be fair cases representing real-world situations in which a member of the public might seek medical information or advice from a symptom assessment app, or present to primary care. Most of the clinical vignettes were newly presenting problems experienced by an individual or by a child in their care, and they included some patients with chronic conditions, e.g. diabetes, hypertension etc. (see **Supplementary Tables 1 - 3**).

The origin of 32.0% of the vignettes (numbers 1 to 64) were anonymised insights from transcripts of real calls made to NHS Direct (a UK national nurse-led telephone next-steps advice/triage service operational until 2014) which had previously been used as part of an NHS Direct benchmarking exercise for recommended outcomes (these were used with full consent of NHS Direct). The remaining 68.0% of the vignettes were created by the vignette creation team (JC, SU and AB), including joint assignment of the most appropriate main diagnosis and differential diagnoses, as a starting point for the vignette gold-standard answers. The vignettes included the sex and age of the patient, previous medical history (including factors such as pregnancy, smoking, high blood pressure, diabetes, other illnesses), the named primary complaint, additional information on the primary complaint and current symptoms, and information to be provided only “if asked” by the tested-GP or symptom assessment app (see **Supplementary Table 3** for example vignettes). Each vignette was created with a majority-decision-based list of gold standard correct conditions including a main-diagnosis and a list of other differential diagnoses (generally between one and four, but length-varying per vignette, as appropriate to the clinical history).

### Vignette review

The vignettes were reviewed externally by a panel of three experienced primary care practitioners, each with more than 20 years primary care experience (see acknowledgements). The role of the review panel was to make changes to improve quality and clarity, and to set the gold-standard main diagnosis and differential diagnoses; this was determined by the majority view.

The gold-standard triage level was set independently of vignette creation, vignette review and vignette diagnosis gold-standard setting - this was done by a separate panel of three experienced primary care practitioners using a tie-breaker panel method based on the matching process set out by [6]. The gold-standard optimal triage was assigned by the panel to a six-point scale (see **Table 1**), independent of the native levels of urgency advice of any of the 8 apps. The tested-GPs’ triage and the levels of urgency advice of each app were mapped to this scale using the linear mapping set out in **Supplementary Fig. 1**.

**Table 1.** Triage levels assigned to each clinical-vignette.

| Level of urgency advice | Description |
| --- | --- |
| 1 | call ambulance |
| 2 | go to emergency department |
| 3 | see primary care within 4 hours |
| 4 | see primary care same day |
| 5 | see primary care non-urgent |
| 6 | home care |

### Assessment of apps and GPs using vignettes

Seven external GPs were tested with the vignettes (the ‘tested-GPs’), providing condition suggestions (preliminary diagnoses) for the clinical vignettes after telephone consultations with JC, who had the role of ‘patient-actor’-physician. All tested-GPs were listed on the GP Register and licensed to practice by the UK General Medical Council and had an average of 11.2 years clinical experience post-qualification as a doctor and 5.3 years post qualification as a GP. Five of the tested-GPs completed telephone consultations for all 200 clinical-vignettes. One GP completed 130 telephone consultations but had to withdraw due to personal reasons. Another GP completed 100 telephone consultations but had to withdraw due to work commitments. Based on the information provided in the telephone consultation, the GPs were asked to provide a main diagnosis, up to five other differential diagnoses, and a single triage level (appropriate to a telephone triage setting).

### Assessment of vignettes by the symptom assessment apps and “coverage”

The clinical vignettes were entered into 8 symptom assessment apps by 8 primary care physicians playing the role of ‘patient’ - (app-entry-Dr-1 to -8 in **Fig. 1**). The versions of the symptom assessment mobile apps assessed were the most up to date version available for iOS download between the dates of 19 Nov. 2019 and 9 Dec. 2019. The version of the Buoy online symptom assessment tool used was the version available online between the dates of 19 Nov. 2019 and 16 Dec. 2019. The symptom assessment apps investigated were Ada, Babylon, Buoy, K Health, Mediktor, Symptomate, WebMD, Your.MD (see **Supplementary Table 4** for a description of these apps). The 8 physicians were listed on the GP Register and licensed to practice by the UK General Medical Council, with at least 2 years of experience as a GP and had never worked or consulted for Ada Health; these physicians had no other role in this study. Each physician entered 50 randomly assigned vignettes (out of 200) into each of four randomly assigned symptom assessment apps. If the app did not allow entry of the clinical vignette (lack of coverage), the reason for this was recorded, as was the reason for every vignette for which condition-suggestions or levels of urgency advice were not provided. If entry was permitted, the physician recorded the symptom assessment app’s condition suggestions and levels of urgency advice and saved screenshots of the app’s results to allow for source data verification. In this way, each vignette was entered once in each app, with four physicians entering vignettes in each app.

### Source data verification

Source data verification was carried out (100% of screenshots compared to spreadsheet data) and any missing or inaccurately transcribed data in the spreadsheets was quantified, recorded in this report and corrected to reflect the screenshot data.

### Metrics for assessing condition-suggestion accuracy

We compared the top-1 suggested condition (M1), the top-3-suggested conditions (M3), and the top-5 suggested conditions (M5) provided by the 7 tested GPs and the 8 apps to the gold-standard main diagnosis. We also calculated the comprehensiveness and relevance of each GP’s and each app’s suggestions [13] - see **Table 2** for a description of the metrics used for comparing condition-suggestion accuracy.

**Table 2.**
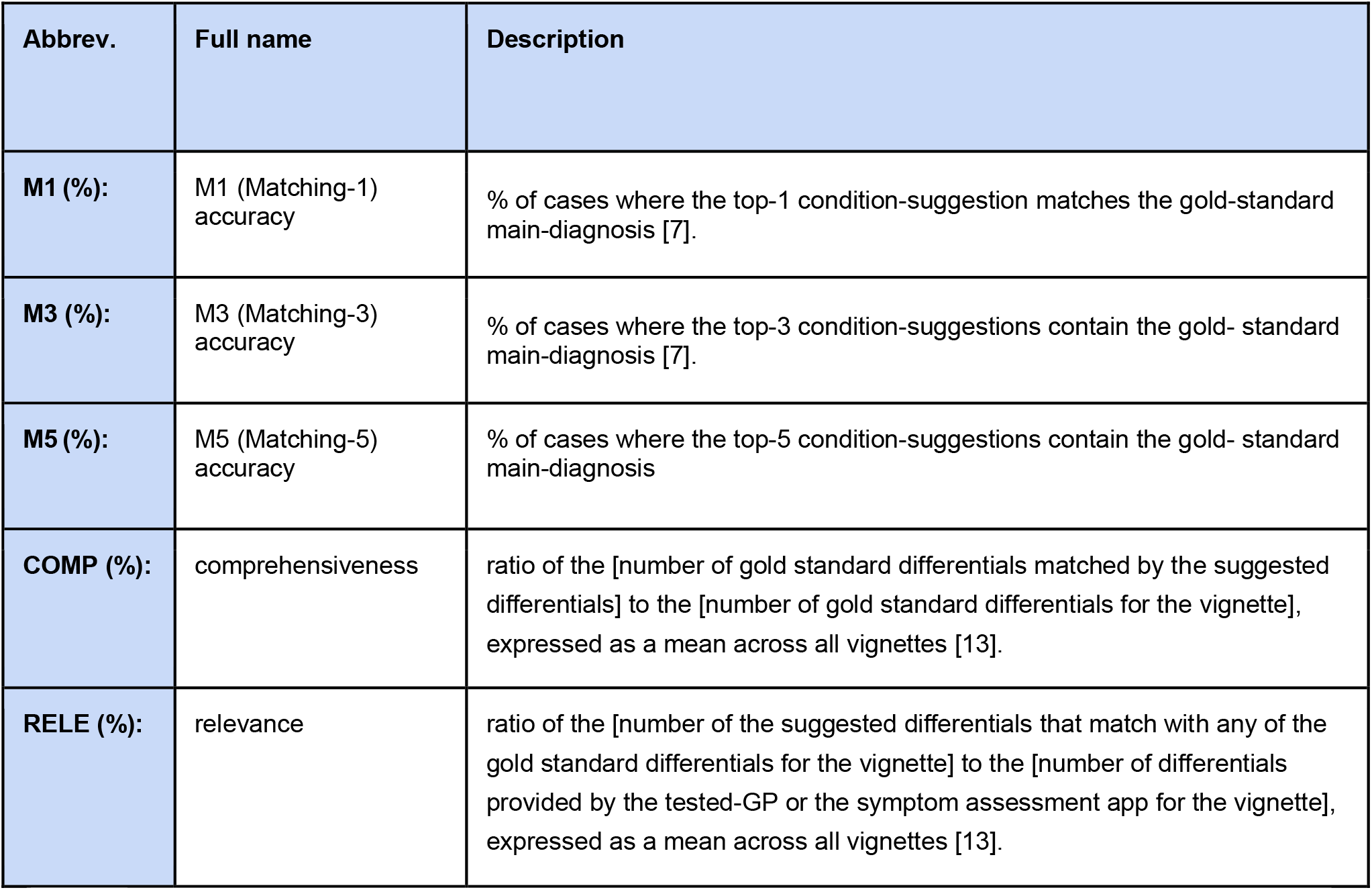
Metrics used in comparison of condition-suggestion accuracy.

| Abbrev. | Full name | Description |
| --- | --- | --- |
| **M1 (%):** | M1 (Matching-1) accuracy | % of cases where the top-1 condition-suggestion matches the gold-standard main-diagnosis [7]. |
| **M3 (%):** | M3 (Matching-3) accuracy | % of cases where the top-3 condition-suggestions contain the gold- standard main-diagnosis [7]. |
| **M5 (%):** | M5 (Matching-5) accuracy | % of cases where the top-5 condition-suggestions contain the gold- standard main-diagnosis |
| **COMP (%):** | comprehensiveness | ratio of the [number of gold standard differentials matched by the suggested differentials] to the [number of gold standard differentials for the vignette], expressed as a mean across all vignettes [13]. |
| **RELE (%):** | relevance | ratio of the [number of the suggested differentials that match with any of the gold standard differentials for the vignette] to the [number of differentials provided by the tested-GP or the symptom assessment app for the vignette], expressed as a mean across all vignettes [13]. |

### Assigning matches between tested-GPs/apps and the gold-standard

Every suggested condition from the tested-GPs and the apps was submitted anonymously to an independent panel of experienced primary care physicians to decide if the suggested condition matched the gold-standard diagnoses list, unless there was an explicit exact match - i.e. identical text of the answer from the tested-GP/app and the gold-standard. Matching was decided using a tie-breaker panel method which was based on the method set out by [6]. The panel was presented with the condition-suggestions blinded to their source. Panellists were instructed to use their own clinical judgement in interpreting whether condition-suggestions were matches to the gold standard, supported by matching criteria (see **Supplementary Table 5**).

#### Mapping and comparing levels of urgency advice

Triage suggestions from each GP and levels of urgency advice from each app were mapped to the gold standard triage-levels using the simple linear mapping scheme set out in **Supplementary Fig. 1**. The degree of deviation of GP triage urgency and of app levels of urgency advice was compared by reporting the percentage of vignettes for which GPs and symptom assessment apps were: (i) overconservative; (ii) overconservative but suitable (one level too high); (iii) exactly-matched; (iv) safe but underconservative (one level too low); or, (v) potentially unsafe.

The WebMD assessment report only provides information on whether each suggested condition is urgent (via an urgency ‘flag’). Finer urgency advice on each condition suggestion is available by clicking through to a separate detailed screen on each suggested condition, but unlike the other apps, no overall vignette-level summary urgency advice is provided. Meaningful comparison to the other apps or tested-GPs was therefore not possible and WebMD was excluded from the urgency advice analysis in this study.

We used confusion matrices in order to fully visualise the severity of misclassification of advice levels [14]. These confusion matrices were weighted in order to represent the relative seriousness of inappropriate urgency advice, either in the direction of being overly conservative (e.g. inefficient use of health care system resources), or in the direction of being insufficiently conservative (potentially unsafe advice). In this weighting ‘less conservative” level of urgency advice assignment was penalized twice as heavily (double absolute distance) than “more conservative” assignment (absolute distance). The weighted confusion matrices were normalised to correct to the number of vignettes for which urgency advice were provided by each app and tested-GP.

### Statistical methods

M1, M3 and M5 performance as well as levels of urgency advice were compared using descriptive statistics and tests appropriate for categorical data. Chi-Square tests were used to test whether the proportion of correct answers from all apps and from all tested-GPs were drawn from the same distribution. In case of a significant difference, two-sided post hoc pairwise Fisher’s Exact Tests [15,16] were used to compare individual app or tested-GP performances. Comprehensiveness and relevance (COMP and RELE) were assessed by Kruskal-Wallis-H-Test (KW-H-Test) applied to all 15 answer datasets (8 apps and 7 tested-GPs), followed by post hoc pairwise testing using the two-sided Dunn test [15], in cases where there was a significant difference on the KW-H-Test. *p*-values were corrected for multiple comparisons using the Benjamini-Hochberg procedure [17] and considered significant if less than 0.05. In figures, error bars for individual app and tested-GP performance represent 95% confidence intervals (CI). These were calculated using the Wilson-Score method for categorical data (M1, M3 and M5) [18] and using the percentile bootstrap method for COMP and RELE [19]. The mean app and tested-GP scores were calculated as arithmetic means of the M1, M3, M5, COMP and RELE performance for each app and each tested-GP, with error bars that represent the standard deviation (S.D.).

### Patient and public involvement

Patients were not involved in setting the research questions, the design, outcome measures or implementation of the study. They were not asked to advise on interpretation or writing up of results. No patients were advised on dissemination of the study or its main results.

## Results

### Source data verification

For vignette cases where the app-entry-Drs made data recording errors, these were corrected to match the source verification data saved in the screenshots. Full sets of screenshots were recorded by 7 of the 8 app-entry-Drs. One app-entry-Dr (#4) did not record all screenshots for K Health, WebMD and for Your.MD and for this reason a subanalysis of the 150 vignettes for which full verification was possible for these apps is provided in **Supplementary Tables 6 & 7**. The differences in performance in this subanalysis is relatively minor and might be due to random differences between the 150 and full vignette sets or be due to app-entry-Dr-4 recording error.

### App coverage

The apps varied in the proportion of vignettes for which they provided any condition-suggestions (see **Fig. 2** and **Supplementary Tables 8 - 10**). The reasons that some apps did not provide condition-suggestions included: (i) not included in the apps’ regulatory “Intended Use” or another product design reason (e.g. users below a set age limit, or pregnant users); (ii) not suggesting conditions for users with severe symptoms (or possible conditions); (iii) presenting complaint not recognised by the app (even after rewording and use of synonyms); and, (iv) some apps did not have coverage for certain medical specialties e.g. mental health. For 12% of the vignettes, the urgency advice from for K Health was not recorded due to app-entry-Dr-4 recording error and was not recorded in source verification data saved in the screenshots. The missing data is labelled in **Fig. 2** and in the later figures describing the appropriateness of urgency advice. A subanalysis of the 150 vignettes for which full data and full verification was possible for K Health is provided in **Supplementary Table 6**.

**Figure 2.**
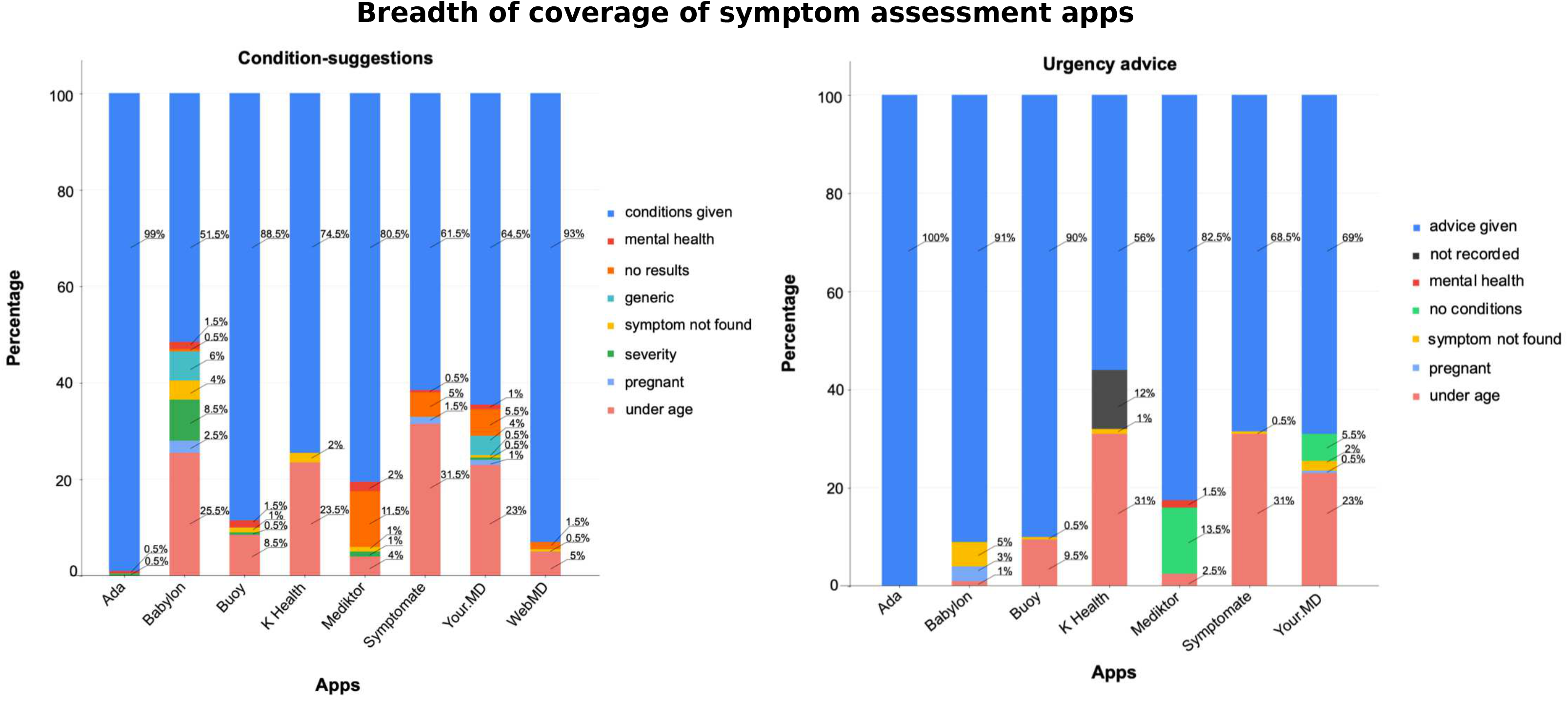
App breadth of coverage - i.e. the proportion of vignettes for which condition-suggestions and levels of urgency advice were provided. When condition-suggestions or urgency were not provided, the principal reason for this is shown, or alternatively ‘no results’ when no reason was given. conditions given - condition-suggestions were provided by the app; mental health - mental health vignettes where no condition-suggestions/urgency advice was provided; no results - the app provided a clear statement that no condition-suggestion results were found for the vignette (the reason why the app failed to give a condition-suggestion for these vignettes is uncertain, but generally these vignettes relate to minor conditions, and in most cases it seems that the app does not have a matching condition modelled); generic - the app gave a generic answer rather than a condition, e.g. “further assessment is needed”; symptom not found - a directly or appropriately matching symptom to the presenting complaint could not be found in the app so the vignette could not be entered; severity - the app did not to give condition-suggestions for very serious symptoms - e.g. the app stated only ‘Condition causing severe [symptom]’; pregnant - vignettes for which no condition-suggestions/urgency advice was provided by the app as the patient was pregnant; under age - vignettes for which no condition-suggestions/urgency advice was provided by the app as the patient was under its specified age limit; advice given - level of urgency advice was provided by the app; not recorded - one app-entry-Dr (#4) did not fully record the levels of urgency advice, and there were no corresponding source data verification screenshots for this subset of data (see **Supplementary Table 6** for a subanalysis of the 150 vignettes with complete source-data-verified data for K Health on levels of urgency advice); no conditions - no condition-suggestions were provided by the app, and, as a result of this, the app did not provide urgency advice. See **Supplementary Tables 8 - 10** for details.

### Suggested conditions: the ‘required-answer’ approach

The approach adopted in other vignettes studies by [3,6,20–24] has been to determine the percentage of all vignettes for which the app (or tested-GP) provided an appropriate condition- suggestion - here this analysis method is referred to as the ‘required-answer’ approach. Results are shown in **Fig. 3**. For a full description for each metric see **Table 2**.

**Figure 3.**
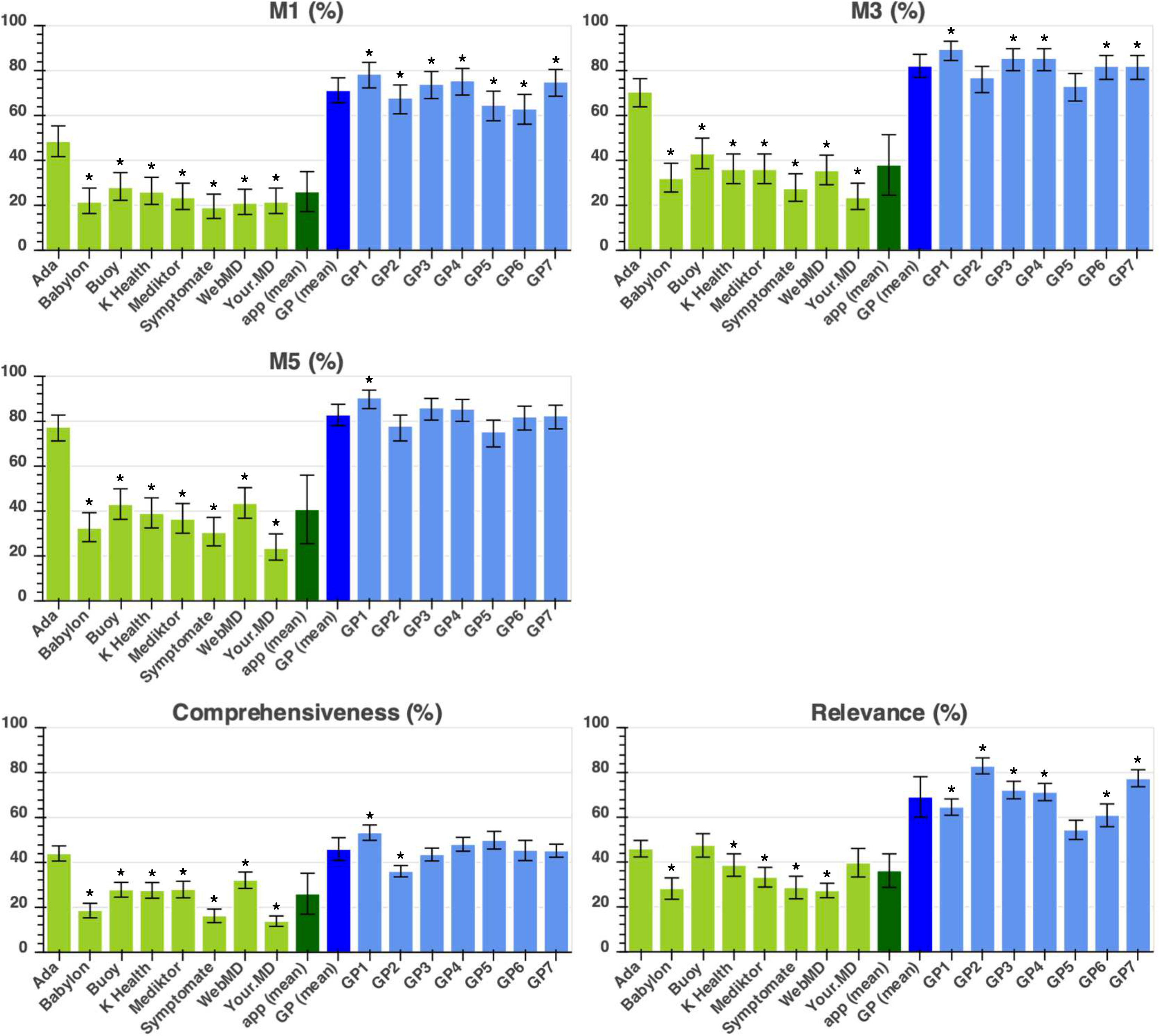
Required-answer approach showing the performance metrics (M1, M3, M5, comprehensiveness and relevance - as defined in **Table 2**) of the 8 apps and 7 tested-GPs. App performance is coloured in light green, average (mean) app performance is in dark green, average (mean) tested-GP performance in dark blue, and individual tested-GP performance in light blue. Statistical significance of the difference between the app with highest performance and all other apps/tested-GPs is shown with the * symbol indicating: *p*-value < 0.05. For one of these apps (Your.MD) one app-entry-Dr (#4) did not record all screenshots needed for source data verification - see **Supplementary Table 7** for a subanalysis of fully verified data, which shows the same trend of results and no significant difference to the data recorded here).

### Suggested conditions: the ‘provided-answer’ approach

For users or physicians choosing or recommending a symptom assessment app, it is relevant to know not only the app accuracy, but also how wide is its coverage and therefore the ‘required-answer’ analysis in the previous section is the most relevant analysis. An alternative approach is the provided-answer analysis, which is the number of correct suggested conditions provided by an app for each vignette *for which it provides an answer*. In other words, there was no penalty for an app that, for any reason, does not provide condition-suggestions for a vignette e.g. children under 2 years old (see **Supplemental Tables 4 & 10**). Both analyses are provided in this study in order to give a fully balanced overview of the performance of all the apps. The results for the provided-answer analysis are shown in **Fig. 4**. For a full description for each metric see **Table 2**.

**Figure 4.**
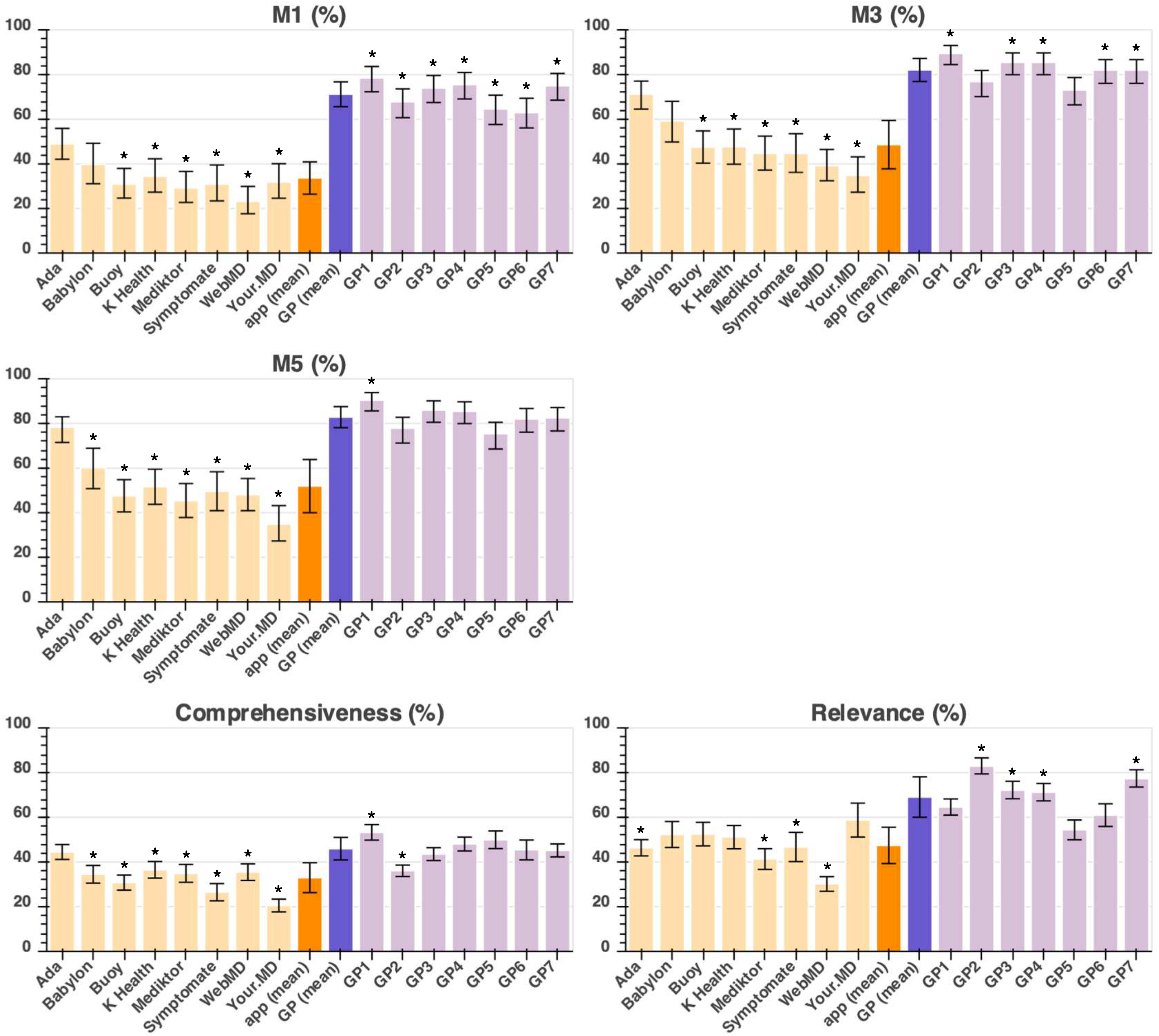
Provided-answer approach showing the performance metrics (M1, M3, M5, comprehensiveness and relevance - as defined in **Table 2**) of the 8 apps and 7 tested-GPs. App performance is coloured in light orange, average (mean) app performance is in dark orange, average (mean) tested-GP performance in dark purple, and individual tested-GP performance in light purple. Statistical significance of the difference between the app with highest performance and all other apps/tested-GPs is shown with the * symbol indicating: *p*-value < 0.05. For one of these apps (Your.MD) one app-entry-Dr (#4) did not record all screenshots needed for source data verification - see **Supplementary Table 7** for a subanalysis of fully verified data, which shows the same trend of results and no significant difference to the data recorded here).

### Levels of urgency advice

The urgency advice performance of each app is summarised in **Table 3**. Tested-GPs had safe triage performance of 97.0±2.5% (where safe is here defined as maximum one level less conservative than gold-standard, expressed per vignette provided with advice) - three apps had safety performance within 1 S.D. of GPs (mean) - Ada: 97.0%; Babylon: 95.1%; and, Symptomate: 97.8%. One app had a safety performance within 2 S.D.s of GPs - Your.MD*: 92.6%. Three apps had a safety performance outside 2 S.D.s of GPs - Buoy: 80.0% (p<0.001); K Health*: 81.3% (p<0.001); Mediktor: 87.3% (p=1.3X10-3) (* - for two of these apps one app-entry-Dr (#4) did not record all screenshots needed for source data verification - see **Supplementary Table 6** for a subanalysis of fully verified data, which shows the same trend of results and no significant difference to the data recorded here).

**Table 3.** Triage levels assigned to each clinical-vignette, where safe is defined as maximum one level less conservative than gold-standard, expressed per vignette provided with advice. * *p*-value < 0.05. For two of these apps (K Health & Your.MD) one app-entry-Dr (#4) did not record all screenshots needed for source data verification - see **Supplementary Table 6** for a subanalysis of fully verified data, which shows the same trend of results and no significant difference to the data recorded here). This analysis is for those vignettes for which urgency advice was provided (i.e. a ‘provided answer) analysis. NS - no significant difference.

| App/<br>tested-GP | Percentage of<br>safe advice | p-value<br>(difference to<br>GP mean) |
| --- | --- | --- |
| Ada | 97.0 | NS |
| Babylon | 95.1 | NS |
| Buoy | 80.0 | <0.001 * |
| K Health | 81.3 | <0.001 * |
| Mediktor | 87.3 | 1.3×10<sup>-3</sup> * |
| Symptomate | 97.8 | NS |
| Your.MD | 92.6 | NS |
| App mean ± S.D. | 90.1±7.4 | - |
| GP mean ± S.D. | 97.0±2.5 | - |
| GP1 | 96.0 | NS |
| GP2 | 96.9 | NS |
| GP3 | 94.0 | NS |
| GP4 | 99.0 | NS |
| GP5 | 100.0 | NS |
| GP6 | 93.9 | NS |
| GP7 | 99.5 | NS |

Fig. 5 summarises and compares urgency advice performance, including the proportion of vignettes for which some apps did not provide advice.

The visualisation in **Fig. 5** provides a high-level overview of urgency advice performance, however a limitation of this approach is that the full range of comparisons between gold standard triage and levels of urgency advice are not shown. The full range of over-conservative and potentially unsafe urgency advice provided by each app and tested GP is shown in the weighted confusion matrices (**Fig. 6**). Low numbers in the matrices (coloured green and yellow) correspond to good urgency advice allocation, high numbers (coloured orange and red) correspond to bad urgency advice allocation. In order to visualise the overall urgency advice performance of each app, i.e. performance both in urgency advice coverage and in the percentage of safe advice, these measures are plotted against each other in **Fig. 7**.

**Figure 5.**
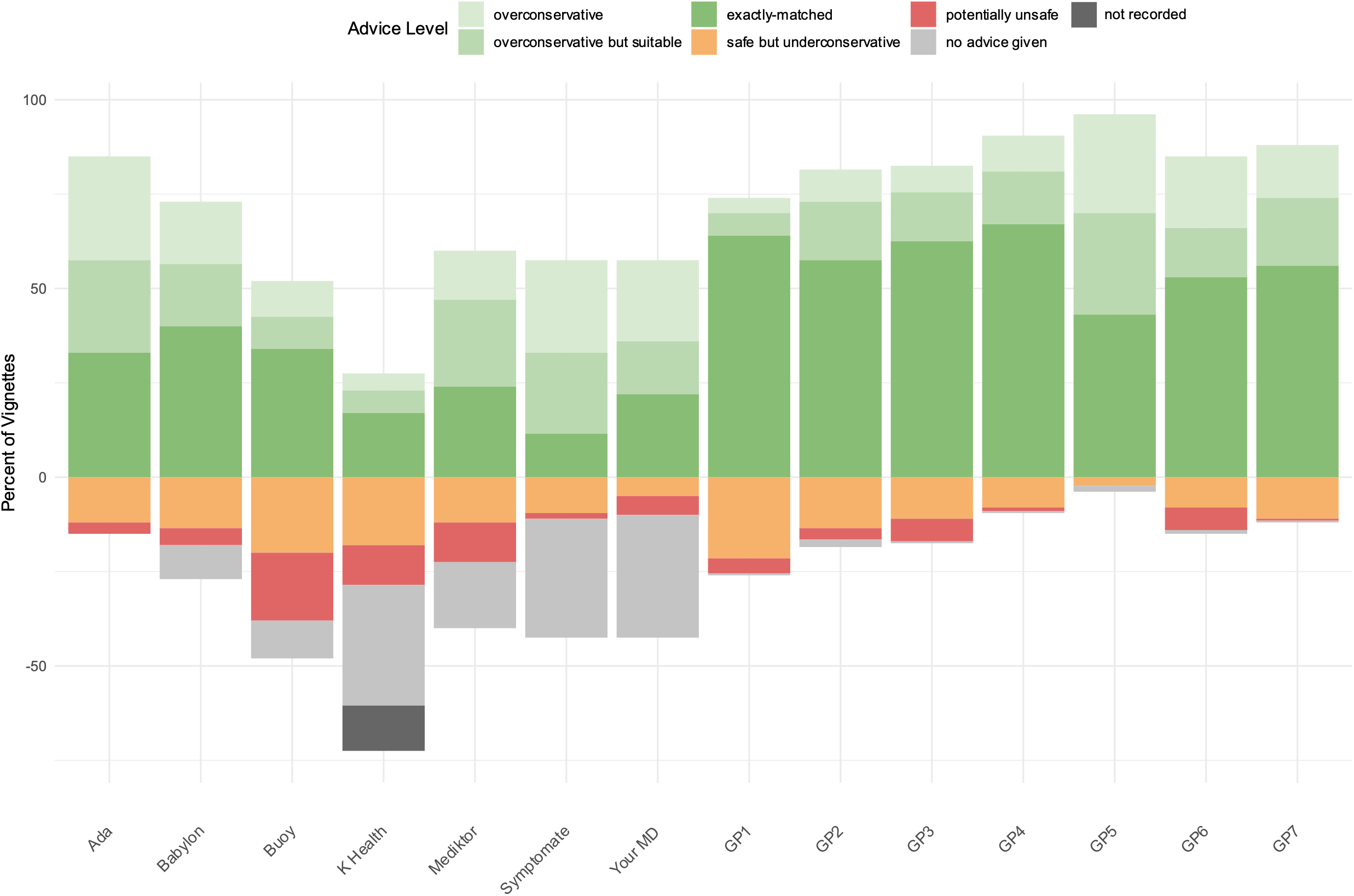
Accuracy of urgency advice displayed as a stacked bar chart centred on the gold standard triage. For two of these apps (K Health & Your.MD) one app-entry-Dr (#4) did not record all screenshots needed for source data verification - see **Supplementary Table 6** for a subanalysis of fully verified data, which shows the same trend of results and no significant difference to the data recorded here).

**Figure 6.**
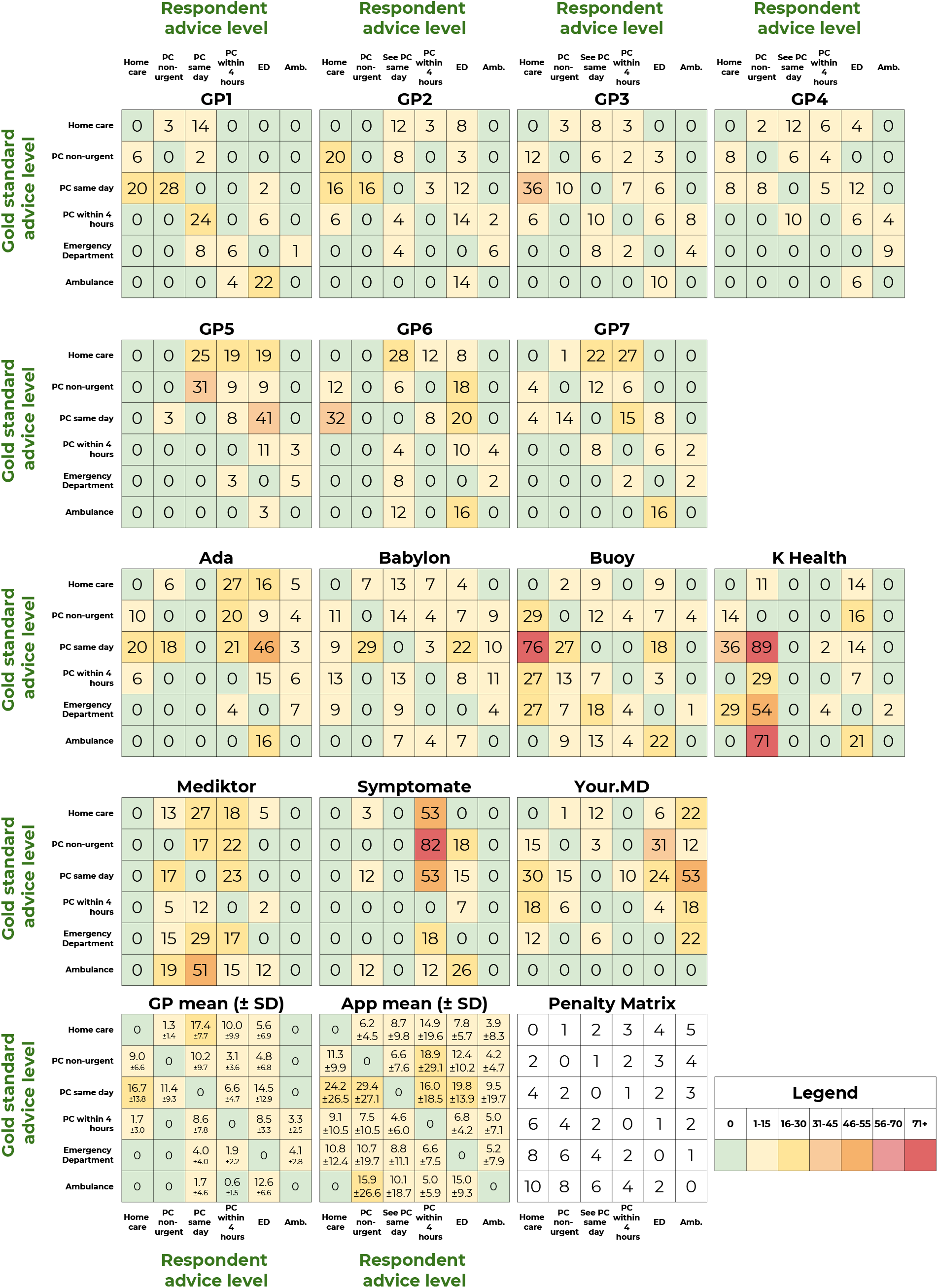
Weighted confusion matrices showing the detailed triage assignments for each app. PC - primary care; ED - emergency department; Amb. - ambulance. For two of these apps (K Health & Your.MD) one app-entry-Dr (#4) did not record all screenshots needed for source data verification - see **Supplementary Table 6** for a subanalysis of fully verified data, which shows the same trend of results and no significant difference to the data recorded here).

**Figure 7.**
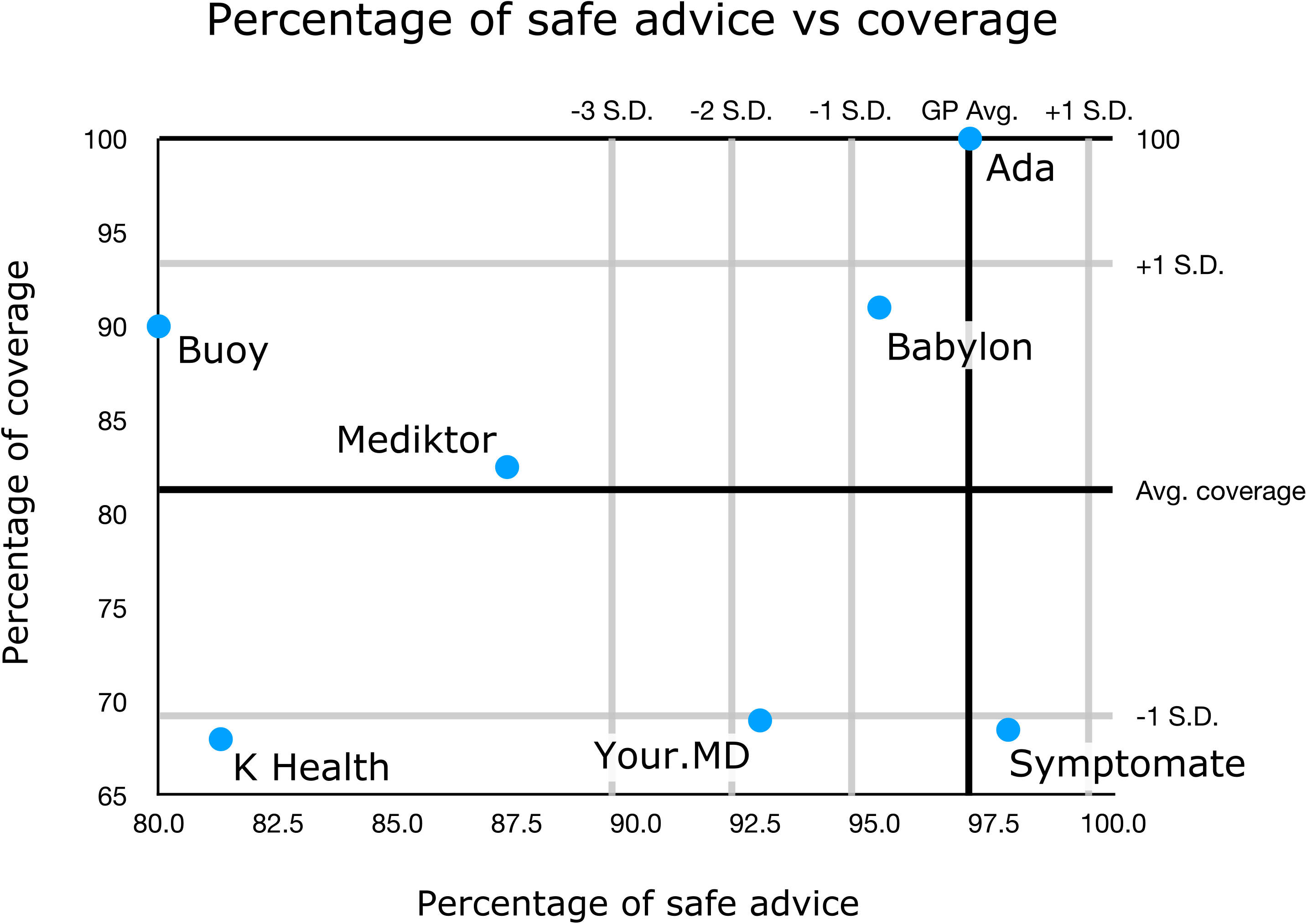
Summary plot of the urgency advice performance of each app. The urgency advice coverage of each app (with respect to app average) is plotted against the percentage of safe advice (with respect to the GP average). For two of these apps (K Health & Your.MD) one app-entry-Dr (#4) did not record all screenshots needed for source data verification - see **Supplementary Table 6** for a subanalysis of fully verified data, which shows the same trend of results and no significant difference to the data recorded here).

## Discussion

### Principal findings

In this clinical vignette comparison of symptom assessment apps and GPs, we found that apps varied substantially in coverage, appropriateness of urgency-advice, and accuracy of suggested conditions.

Synthesising the analyses on the appropriateness of urgency advice (see **Table 3** and **Fig. 5 - 7**), the apps can be categorised as follows:

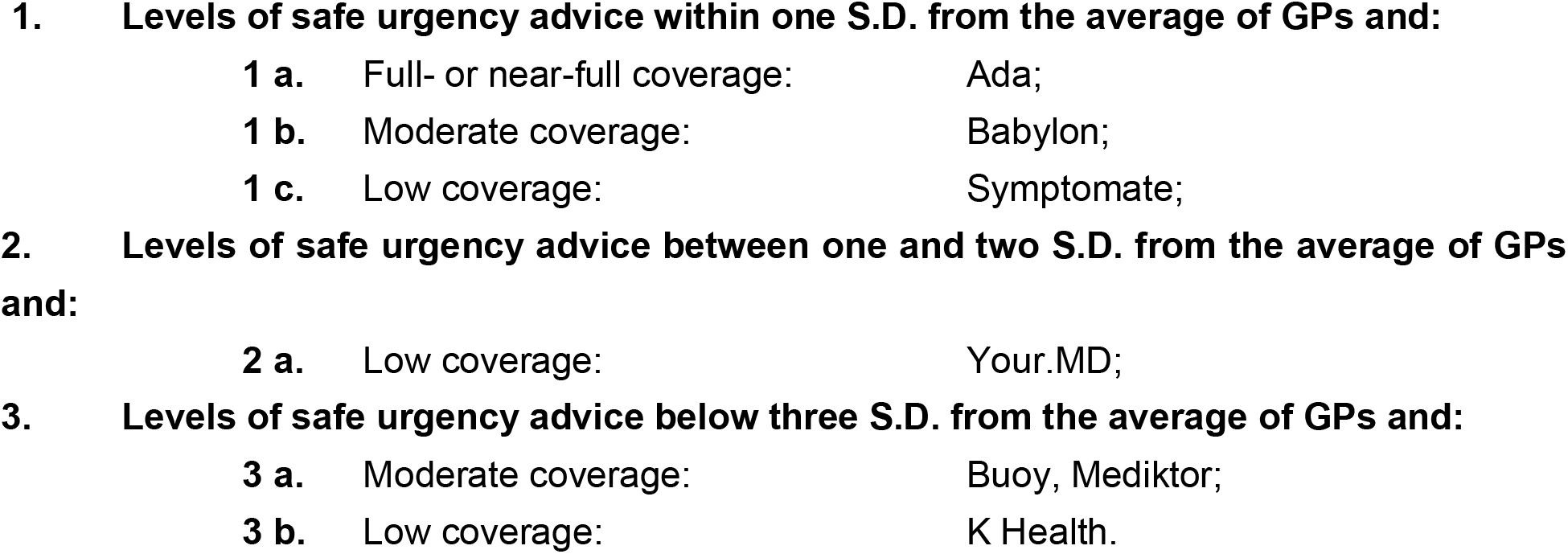

Condition-suggestion coverage varies greatly with a range of 47.5% from highest (Ada; 99.0%) to lowest (Babylon, 51.5%). Although there is no absolute cut-off of what an acceptable condition suggestion coverage is, an app that can provide high coverage along with a high accuracy of condition-suggestion and high urgency advice appropriateness, will generally be superior to an app with narrow coverage. There is no identifiable correlation between app M1 or M3 condition-suggestion accuracy or urgency-advice accuracy and the condition-suggestion coverage or urgency-advice coverage.

There was considerable variation in condition-suggestion accuracy between the GPs and between apps. For top-1 condition-suggestion (M1), the range of tested-GPs was 16.0%, the S.D. 5.6% and for M3 the range was 15.9% and S.D. 5.2%; For the apps, the M1 condition-suggestion accuracy range was 29.5%, the S.D. 8.9% and the M3 range was 47.0% and S.D. 13.5%. The GPs all outperformed apps for top-1 condition matching. For M3 and M5 (i.e. including the gold standard diagnosis in top-3 and -5 suggestions), the best performing app (Ada) was comparable to tested-GPs, with no significant difference between its performance and the performance of several of the tested-GPs. The top performing symptom assessment app (Ada) had an M3 27.5% percent higher than the next best performing app (Buoy, *p*<0.001) and 47.0% higher than the worst-performing app (Your.MD, *p*<0.001). There was a significant difference between top performing app (Ada) and other apps for all condition accuracy measures, with two exceptions for relevance (in the required-answer analysis).

There was also considerable variation in urgency advice performance between the GPs and between apps. The range of tested-GP safe advice was 6.0% and the S.D. was 2.5%; for the apps the range of safe advice was 17.8% and the S.D. 7.4%. Tested GPs had an average safe advice performance of 97.1±2.5% - only three apps had safe advice performance within 1 S.D. of the GPs (mean) - Ada: 97.0%; Babylon: 95.1%; and, Symptomate: 97.8%.

### Strengths and limitations of this study

The systematic review of Chambers et al. [4] identified limitations of published studies on the safety and accuracy of symptom assessment apps as: (i) not being based on real patient data; (ii) not describing differences in outcomes between symptom assessment apps and health professionals; (iii) covering only a limited range of conditions; (iv) covering only uncomplicated vignettes; and, (v) sampling a young healthy population not representative of the general population of users of the urgent care system. Of these limitations, only one applies to this study - the limitation of being based on clinical-vignettes rather than on real-patient data. The effect of this limitation has been minimised through the development of many of the vignettes to be highly realistic through the use of anonymised real-patient data collated from NHS Direct transcripts. The use of real patient data with an actual diagnosis is not without its limitations in the evaluation of symptom assessment app accuracy as it relies on face-to-face consultation to confirm diagnosis. Very often diagnosis is only provided after physical examination or diagnostic tests, so comparison is confounded as the real patient diagnosis is based on additional information not made available to the app. The vignettes approach has allowed this study to be designed to minimise the limitations (ii) to (v) identified by Chambers et al. [4].This has been done for limitation (ii) through inclusion of a 7-GP comparator group; for limitation (iii) by development of vignettes for conditions spanning all body systems and sampling all medical specialisms relevant to primary care presentation; for limitation (iv) by designing clinical-vignettes including not-only simple and common situations, but also moderately complex and challenging presentations; for limitation (v) through including vignettes spanning from 1-month to 89-years old.

Relative strengths of this study are the large number of clinical-vignettes included (n=200), along with the separation in the design of clinical-vignette writing from the process of deciding on the gold-standard main and secondary differential-diagnoses and appropriate levels of urgency advice. Another strength of this study is that GPs were tested with vignettes in a manner that simulates real clinical consulting - in this way the GPs consultation process was assessed, enabling a fair comparison to the apps. Vignettes were entered into the apps by 8 additional primary care physicians acting as the user (app-entry-Dr-1 - 8). A physician also ‘acted’ as the patient being assessed by the GPs in the phone-consultations. It has been argued that lay-person entry is closest to the real intended use of symptom assessment apps [25], however, it is known that lay-people are less reliable at entering clinical-vignettes than health care providers [26]. A further strength of this study is that each decision of whether a condition-suggestion (from an app or a GP) matched the clinical-vignette’s main and other differential-diagnoses was made in a rigorous manner following the 3-physician tie-breaker panel approach of [6].

A limitation of this study was that a systematic and comprehensive selection process was not used to select the symptom assessment apps to be included. The non-systematic selection criteria used were that, at the time of selection: (i) Babylon and Ada are leading symptom assessment smartphone apps in the UK; (ii) K Health, WebMD and Ada are the most used in US (usage data from Sensely, https://www.sensely.com); (iii) Mediktor and Buoy have existing published data [7,27]; and, (iv) Your.MD has a similar user experience and user interface to Babylon and Ada and has been compared with them in small non-peer reviewed studies [21,23].

Direct comparison of levels of urgency advice between individual apps and between apps and GPs was challenging because i) some apps provided no levels of urgency advice for large numbers of vignettes; (ii) performing well in one level of urgency advice trades off performance in other levels of urgency advice; (iii) the nature of urgency advice reporting was different in WebMD (see Methods section).

Furthermore, the vignettes may have had a UK bias and some of the symptom assessment apps (e.g. Buoy, K Health & WebMD) are primarily used in the US. A data acquisition error by one of the app-entry-Drs meant there were unrecorded urgency advice data for 12.0% of vignettes for one app (K Health) and incomplete source data verification screenshots for two other apps (Your.MD and WebMD). The implication for this for the main analysis was investigated in two subanalyses in the data supplement. Future studies could ensure ongoing source data verification rather than waiting until the end of study data collection for review. It is an unavoidable limitation that software evolves rapidly, and the performance of these apps may have changed significantly (for better or worse) since the time of data collection. Finally, this study was designed, conducted, and disseminated by a team that includes employees of Ada Health; future research by independent researchers should seek to replicate these findings and/or develop methods to continually test symptom assessment apps.

### Comparisons to the wider literature

The results of this study are qualitatively broadly similar to reported results from other inter-app relative performance studies, including one peer-reviewed study [24] and two non-peer-reviewed studies [21,23]. A peer-reviewed study using 45 Ear, Nose and Throat (ENT) -vignettes [24] evaluated M1 and M3 results and found that Ada had substantially better performance than other apps. Overall, Ada was the second-best performing app out of 24 tested apps in the ENT-discipline [24].

A small non-peer-reviewed independent clinical vignettes study tested NHS 111, Babylon, Ada and Your.MD and found similar overall results to this study [23]; they also found that all apps were successful at spotting serious conditions, such as a heart attack, and that they were fast and easy to use. A second small 2017 non-peer-reviewed independent vignettes study [21], that was carried out by established symptom assessment app academic researchers, tested Babylon, Ada and Your.MD. The trend of the results was similar to those in this study.

In an observational study carried out in a Spanish Emergency Department (ED) waiting room the Mediktor symptom assessment app was used for non-urgent emergency cases for patients above 18 years old [7]. The study calculated accuracy with consideration only for those patients whose discharge diagnosis was modelled by the app at the time. For a total of 622 cases Mediktor’s M1 score was reported as 42.9%, M3 score as 75.4% (i.e. the symptom assessment app’s top-1 (M1), top-3 (M3), or top-10 condition-suggestion(s) matched the discharge-diagnosis in this percentage of cases). When the [7] reported results are refactored to consider all patient discharge-diagnoses (the standard approach) the: M1 is 34.0% and M3 is 63.0%, compared to M1 of 23.5% and M3 of 36.0% for Mediktor in this study (all-vignettes data). The reason for lower Mediktor performance in the current study compared to [7] is not known but it may be related to a different range of conditions or difficulty level than the non-urgent emergency cases presenting to the ED - for example - the vignettes in this study contain many true emergency cases and also many GP or pharmacy/treat-at-home cases which would not be represented by the ED patients included in [7]. In a 2017 42-vignette evaluation of WebMD, [28] determined its accuracy for ophthalmic condition suggestion: M1 was 26.0% and M3 was 38.0%. Urgency advice based on the top diagnosis was appropriate in 39.0% of emergency cases and 88.0% non-emergency cases.

The manufacturers of the apps Babylon and Your.MD responded to the two non-peer reviewed studies [21,23] observing that their apps have been updated and improved subsequent to the publication of those reports. Nevertheless, the findings with respect to condition-suggestion performance, in the later peer-reviewed study by Nateqi et al. [24] and in the present study appear to be in line with those from the two non-peer reviewed studies.

### Implications for clinicians and policymakers

The results of this study are relevant for home users of symptom assessment apps, and to health care providers offering advice to their patients on which symptom assessment apps to choose. There are large (and statistically significant) differences between app coverage, suggested condition accuracy and urgency-advice accuracy. One of the biggest challenges in comparing symptom checker apps are the differences in coverage. Some coverage restrictions, such as not allowing symptom assessment for one user subgroup (e.g. children), have no negative effect on the app’s effective use for other user subgroups (e.g. adults). Other situations, such as the inability to search for certain symptoms, providing no condition-suggestions / urgency advice for certain input symptoms, or, excluding co-morbidities, mental health or pregnancy are more problematic and can raise concerns about the safety and benefits of the app for users who might be in those groups.

### Unanswered questions and future research

Future research should evaluate the performance of the apps compared with real-patient data - multiple separate single-app studies are a very unreliable way to determine the true level of the state of the art of symptom-assessment apps. A positive step in this direction is the ITU/WHO Focus Group AI for Health (FG-AI4H) through which several manufacturers of symptom assessment apps evaluated in this study are working collaboratively to create standardised app benchmarking with independently curated and globally representative datasets (“Focus Group on ‘Artificial Intelligence for Health,’” n.d.)[29]. Additional areas that could be explored in such studies are comparative economic impact, understanding user behaviour following an assessment, i.e. compliance with urgency advice (extending the approach of [27]) and impact on health services usage, and, the impact of using the apps to complement a standard GP consult (e.g. through diagnostic-decision support). While it has been argued that the accuracy of urgency advice may be the most important output from a health assessment app, the condition suggestions may be valuable to support patient decision making [27]. To address the effect of patients entering data directly into an app about their own acute conditions, an observational investigation is currently underway in an acute clinical setting in the US by investigators including co-authors of this study. This includes a survey of users’ technological literacy and user experience.

## Conclusions

This study provides useful insights into the relative performance of 8 symptom-assessment apps, compared to each other and compared to 7 tested-GPs, in terms of their coverage, their suggested condition accuracy and the accuracy of their levels of urgency advice. The results show that the best performing of these apps have a high level of urgency advice accuracy which is close to that of GPs. Although not yet as accurate as GPs in top-1 suggestion of conditions, the best apps are close to GP-performance in providing the correct condition in their top-3 and top-5 condition-suggestions.

While no digital tool out-performed GPs in this analysis, some came close, and the nature of iterative improvements to software suggests that further improvements will occur with experience and additional evaluation studies.

## Data Availability

All data relevant to the study are included in the article or uploaded as supplementary information with the exception of the case vignettes, which will not be uploaded because they will be used in periodic update of the study analysis (in order to monitor comparatively change in app performance over time). Publication would prevent this important ongoing scientific research. The vignettes will not be disclosed to the Ada medical intelligence team or to other app developers.

## Footnotes

- Author Statement SG, AM, CC, JC, HF, FP, ET, VV, NV & CN contributed to the planning (study conception, protocol development). SG, AM, AB, CC, JC, FP, CR, SU, VV, NV, & CN contributed to the conduct (coordination of vignette creation, review, coordination of GP or app-testing, condition-matching panel coordination). SG, AM, CC, JC, HF, EM, JM, FP, CR, ET, NV, PW & CN contributed to the data analysis & interpretation. SG, AM, JC, HF, JM, FP, CR, ET, VV, NV, PW & CN contributed to the reporting (report writing). All the authors contributed to commenting on drafts of the report. SG is the guarantor for this work. The corresponding author attests that all listed authors meet authorship criteria and that no others meeting the criteria have been omitted.
- **Disclosures** SG, AM, AB, CC, EM, JM, FP, CR, ET, SU, VV, NV & CN are employees or company directors of Ada Health GmbH and some of the listed hold stock options in the company. JC and PW have or have had consultancy contracts with Ada Health GmbH. The Ada Health GmbH research team has received research grant funding from Fondation Botnar and the Bill & Melinda Gates Foundation. PW has received speaker fees from Bayer and honoraria from Roche, ARISLA, AMIA, IMI, PSI, and the BMJ.
- **Funding** This study was funded by Ada Health GmbH. HF has not received any compensation from Ada Health financial or otherwise.
- **Disclaimer** None
- **Competing interests** Some of the authors are employees of/hold equity in the manufacturer of one of the tested apps (Ada Health GmbH). See author affiliations.
- **Patient consent for publication** Not relevant - no direct patient data.
- **Provenance and peer review** Not commissioned; externally peer reviewed.
- **Data availability statement** All data relevant to the study are included in the article or uploaded as supplementary information with the exception of the case vignettes, which will not be uploaded because they will be used in periodic update of the study analysis (in order to monitor comparatively change in app performance over time). Publication would prevent this important ongoing scientific research. The vignettes will not be disclosed to the Ada medical intelligence team or to other app developers.

## Acknowledgments

Paul Taylor (UCL Institute of Health Informatics) independently reviewed and made suggestions on the study protocol, and after study data collection was complete, reviewed and made suggestions in a draft of this manuscript with respect to the analysis approach and the study description. Vignette review was carried out by the following experienced primary care physicians: Alison Grey, Helen Whitworth, Jo Leahy. Study support was provided by Linda Cen (Data Engineer, Ada Health GmbH), Leif Ahlgrimm (Student IT System Administrator, Ada Health GmbH), Neil Rooney (Senior IT System Administrator, Ada Health GmbH).

## References

1 Mcdaid D, Park A-L. Online Health: Untangling the Web. 2011.

2 Van Riel N, Auwerx K, Debbaut P, et al. The effect of Dr Google on doctor-patient encounters in primary care: a quantitative, observational, cross-sectional study. BJGP open 2017;1.

3 Semigran HL, Levine DM, Nundy S, et al. Comparison of Physician and Computer Diagnostic Accuracy. JAMA Intern Med 2016;176:1860–1. doi:10.1001/jamainternmed.2016.6001

4 Chambers D, Cantrell AJ, Johnson M, et al. Digital and online symptom checkers and health assessment/triage services for urgent health problems: systematic review. BMJ Open 2019;9:e027743. doi:10.1136/bmjopen-2018-027743

5 Millenson ML, Baldwin JL, Zipperer L, et al. Beyond Dr. Google: the evidence on consumer-facing digital tools for diagnosis. Diagnosis 2018;5:95–105. doi:10.1515/dx-2018-0009

6 Semigran HL, Linder JA, Gidengil C, et al. Evaluation of symptom checkers for self diagnosis and triage: audit study. BMJ 2015;351:h3480. doi:10.1136/bmj.h3480

7 Moreno Barriga E, Pueyo Ferrer I, Sánchez Sánchez M, et al. [A new artificial intelligence tool for assessing symptoms in patients seeking emergency department care: the Mediktor application]. Emergencias 2017;29:391–6.

8 Greenhalgh T, Wherton J, Shaw S, et al. Video consultations for covid-19. BMJ 2020;368:m998. doi:10.1136/bmj.m998

9 Heymann DL, Shindo N, WHO Scientific and Technical Advisory Group for Infectious Hazards. COVID-19: what is next for public health? Lancet 2020;395:542–5. doi:10.1016/S0140-6736(20)30374-3

10 Converse L, Barrett K, Rich E, et al. Methods of Observing Variations in Physicians’ Decisions: The Opportunities of Clinical Vignettes. J GEN INTERN MED 2015;30:586–94. doi:10.1007/s11606-015-3365-8

11 Evans SC, Roberts MC, Keeley JW, et al. Vignette methodologies for studying clinicians’ decision-making: Validity, utility, and application in ICD-11 field studies. International Journal of Clinical and Health Psychology 2015;15:160–70. doi:10.1016/j.ijchp.2014.12.001

12 Veloski J, Tai S, Evans AS, et al. Clinical Vignette-Based Surveys: A Tool for Assessing Physician Practice Variation: American Journal of Medical Quality Published Online First: 3 September 2016. doi:10.1177/1062860605274520

13 Berner ES, Webster GD, Shugerman AA, et al. Performance of Four Computer-Based Diagnostic Systems. New England Journal of Medicine 1994;330:1792–6. doi:10.1056/NEJM199406233302506

14 Swaminathan S, Qirko K, Smith T, et al. A machine learning approach to triaging patients with chronic obstructive pulmonary disease. PLoS ONE 2017;12:e0188532. doi:10.1371/journal.pone.0188532

15 Nayak BK, Hazra A. How to choose the right statistical test? Indian J Ophthalmol 2011;59:85–6. doi:10.4103/0301-4738.77005

16 Shan G, Gerstenberger S. Fisher’s exact approach for post hoc analysis of a chi-squared test. PLOS ONE 2017;12:e0188709. doi:10.1371/journal.pone.0188709

17 Benjamini Y, Hochberg Y. Controlling the False Discovery Rate: A Practical and Powerful Approach to Multiple Testing. Journal of the Royal Statistical Society Series B (Methodological) 1995;57:289–300. https://www.jstor.org/stable/2346101 (accessed 20 Apr 2020).

18 Wilson EB. Probable Inference, the Law of Succession, and Statistical Inference. Journal of the American Statistical Association 1927;22:209–12. doi:10.1080/01621459.1927.10502953

19 Efron B, Tibshirani, R.J. An introduction to the Bootstrap (Monographs on Statistics and Applied Probability). New York, NY:: Chapman and Hall/CRC 1998.

20 Bisson LJ, Komm JT, Bernas GA, et al. How Accurate Are Patients at Diagnosing the Cause of Their Knee Pain With the Help of a Web-based Symptom Checker?: Orthopaedic Journal of Sports Medicine Published Online First: 19 February 2016. doi:10.1177/2325967116630286

21 Burgess M. Can you really trust the medical apps on your phone? Wired UK 2017. https://www.wired.co.uk/article/health-apps-test-ada-yourmd-babylon-accuracy (accessed 25 Mar 2020).

22 Powley L, McIlroy G, Simons G, et al. Are online symptoms checkers useful for patients with inflammatory arthritis? BMC Musculoskeletal Disorders 2016;17:362. doi:10.1186/s12891-016-1189-2

23 What happened when Pulse tested symptom checker apps. Pulse Today. http://www.pulsetoday.co.uk/news/analysis/what-happened-when-pulse-tested-symptom-checker-apps/20039333.article (accessed 25 Mar 2020).

24 Nateqi J, Lin S, Krobath H, et al. Vom Symptom zur Diagnose—Tauglichkeit von Symptom-Checkern. HNO 2019;67:334–42. doi:10.1007/s00106-019-0666-y

25 Fraser H, Coiera E, Wong D. Safety of patient-facing digital symptom checkers. The Lancet 2018;392:2263–4.

26 Jungmann SM, Klan T, Kuhn S, et al. Accuracy of a Chatbot (Ada) in the Diagnosis of Mental Disorders: Comparative Case Study With Lay and Expert Users. JMIR Form Res 2019;3:e13863. doi:10.2196/13863

27 Winn AN, Somai M, Fergestrom N, et al. Association of Use of Online Symptom Checkers With Patients’ Plans for Seeking Care. JAMA Netw Open 2019;2:e1918561. doi:10.1001/jamanetworkopen.2019.18561

28 Shen C, Nguyen M, Gregor A, et al. Accuracy of a Popular Online Symptom Checker for Ophthalmic Diagnoses. JAMA Ophthalmol 2019;137:690–2. doi:10.1001/jamaophthalmol.2019.0571

29 Wiegand T, Krishnamurthy R, Kuglitsch M, et al. WHO and ITU establish benchmarking process for artificial intelligence in health. Lancet 2019;394:9–11. doi:10.1016/S0140-6736(19)30762-7

